# Pericapsular nerve group block for hip fracture is safe and effective in the emergency department: a prospective observational study comparing pericapsular nerve group block to fascia iliaca compartment block and femoral nerve block

**DOI:** 10.1101/2021.08.31.21262750

**Authors:** Alan Fahey, Elinor Cripps, Aloysius Ng, Amy Sweeny, Peter J. Snelling

## Abstract

**Background:** The pericapsular nerve group (PENG) block was first described for the treatment of hip fracture, including neck of femur, in 2018. We hypothesise that the PENG block is safe and effective for patients with hip fracture when provided by emergency physicians and trainees in the emergency department (ED), for which it may be superior to fascia iliaca compartment block (FICB) and femoral nerve block (FNB).

**Methods:** From October 2019 to July 2020, consecutive patients receiving regional anaesthesia for hip fracture in the ED of a single large regional hospital were prospectively enrolled. Pain scores were assessed prior to regional anaesthesia then at 15, 30 and 60 minutes after regional anaesthesia. Maximal reduction in pain scores within 60 minutes were assessed using the Visual Analogue Scale (at rest and on movement) or the Pain Assessment IN Advanced Dementia tool (at rest). Patients were followed for opioid use for 12 hours after regional anaesthesia and adverse events over the duration of their admission.

**Results:** There were 67 eligible patients during the enrolment period, with 52 (78%) prospectively enrolled. Thirty-three received femoral blocks (19 FICB, 14 FNB) and 19 received a PENG block. There was no difference in maximum pain score reduction between the groups whether measured at rest or on movement. Clinicians providing the PENG block were less experienced in the technique than those providing FICB or FNB. There was no difference in adverse effects between groups. Although opioid use was similar between the groups, more patients were opioid free after a PENG block.

**Conclusions:** Although there was no difference in maximal pain score reduction, this study demonstrated that the PENG block was feasible and could be provided safely and effectively to patients with hip fracture in the ED. On this basis, a larger randomised controlled study should now be designed.

**Key Messages:** **What is already known on this subject**

□ There is a solid neuroanatomical rationale to suggest the PENG block may provide superior anaesthesia of hip fractures than FNB or FICB.
□ The technique utilises bony sonographic and tactile landmarks which make it an ideal block for emergency physicians to safely and effectively perform.
□ **What this study adds**
□ This is the first comparative study of the PENG block to FNB or FICB in patients with hip fracture in ED, which will provide a scaffold for future research.
□ This pragmatic observation of evolving practice showed that emergency physicians and trainees inexperienced in the technique could provide it safely and effectively in the ED

## INTRODUCTION

Proximal femur fractures (including neck of femur fracture) simply referred to as hip fractures, are a common, painful condition of patients presenting to the emergency department (ED) who are typically elderly and frail.^1,2^ Hip fractures are associated with significant morbidity and mortality and the disease burden is expected to increase.^1-2^ ED management of these patients should incorporate multimodal analgesia, including regional anaesthesia, which is a well established standard of care.^3^ Effective regional anaesthesia improves comfort and reduces opioid requirement, which limits opioid related harm.^4^

The fascia iliaca compartment block (FICB) and femoral nerve block (FNB) are both widely accepted as the current standard of care for regional anaesthesia of hip fractures in the ED.^3^ Both of these blocks provide femoral nerve blockade for anaesthesia of the femur but also cause lower limb weakness.^5-6^ There is limited evidence that one is superior in effectiveness to the other.^7,8^

The pericapsular nerve group (PENG) block (Figure 1) is a recently described regional anaesthetic technique that may be superior to both the FICB and FNB for elderly patients with hip fractures, given the potential for more complete anaesthesia of the joint capsule with motor sparing effects.^9,10^ The technique utilises sonographic and tactile bony landmarks to inject local anaesthetic into the iliopsoas plane at a site distant from blood vessels. The sensory branches of the femoral nerve (FN) and accessory obturator nerve (AON) that innervate the anterior hip capsule traverse this potential space at the level of the superior pubic ramus and are anaesthetised by the PENG block.^11,12^ In contrast, effective femoral blockade by the FICB or FNB relies on cranial spread of anaesthetic to block proximal branches of the FN which then travel caudally within the iliacus muscle via the iliopsoas plane to the anterior hip capsule.^12^ These femoral blocks do not block the AON, which is present in up to 54% of patients.^12^ The anterior hip capsule is also innervated by the obturator nerve (ON) in 83-98% of patients^12^ which is not blocked the FICB or FNB^13^ but may be variably anaesthetised by the PENG block.^14^ However, existing evidence for use of the PENG block for analgesia of hip fractures is currently limited.^15^

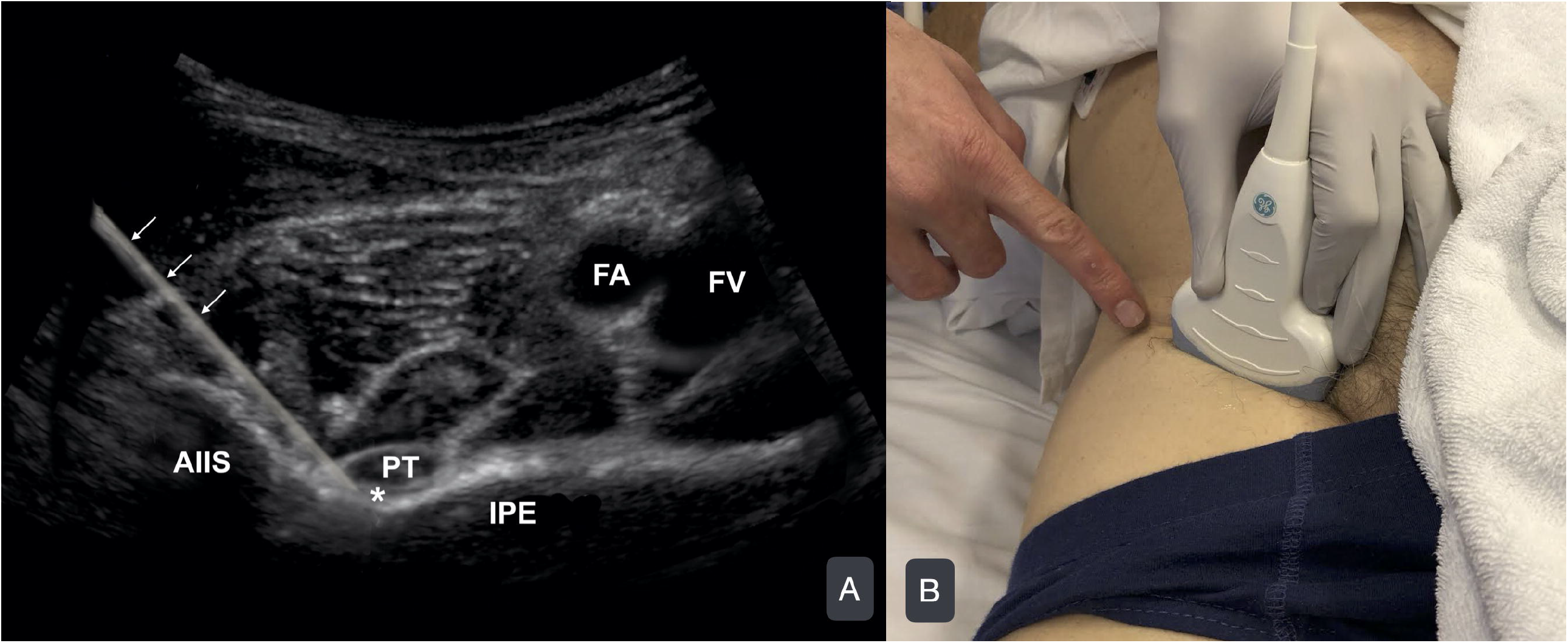

The objective of this study was confirm the feasibility of the PENG block in everyday ED practice and to compare the safety and effectiveness of the PENG block against the current standard regional anaesthesia techniques, FICB and FNB, hereafter combined and referred to as femoral blocks (FB), for patients with hip fracture in the ED. We hypothesise that it is safe and effective for emergency medicine clinicians of variable experience to perform the PENG block for patients presenting to ED with a hip fracture. The primary outcome was the maximal reduction in pain score (VAS, PAINAID) assessed at rest within 60 minutes of the PENG block being administered.

## METHODS

### Study design and setting

This was a single centre prospective, pragmatic, observational cohort study conducted between October 2019 and July 2020 in the ED of a large regional centre in northern New South Wales, Australia. This regional hospital had an ED census of around 33,000 adult patients in 2019. The study was endorsed by emergency and anaesthetic specialists working at the hospital. The study was approved by the North Coast New South Wales Human Research and Ethics Committee and registered in the Australian New Zealand Clinical Trials Registry before commencement (ACTRN12619001410145). We adhered to the STrengthening the Reporting of OBservational studies in Epidemiology statement (https://strobe-statement.org/).

### Selection of participants

Patients were considered for the study if they had a suspected proximal femur fracture. To be eligible for inclusion, a proximal femur fracture, including intracapsular (neck of femur) or extracapsular (intertrochanteric or peritrochanteric), had to be confirmed on either radiograph or computed-tomography (CT) reported by a radiologist, with regional anaesthesia, either FICB, FNB or PENG block, performed by an emergency physician or trainee. Additionally, patients had to be prospectively enrolled with pain scores recorded on a designated clinical research form (CRF). Exclusion criteria included age younger than 18 years, femoral shaft fracture, and regional anaesthesia provided at another location. Patients were screened 24 hours a day, 7 days a week to ensure consecutive recruitment. Participant consent was waived for this observational study of routine practice. Clinicians were not masked to the study objectives.

### Patient and public involvement

There was no public or patient involvement in the study design. However, there was no treatment allocation as part of the study and a patient centred outcome (analgesia) was the primary outcome.

### Interventions

Patients were treated at the discretion of their treating clinician without influence from the investigators. The experience of the clinician performing the regional anaesthesia technique was prospectively documented. All blocks were performed using ultrasound guidance and ropivacaine 75-150mg. Lignocaine (up to 100mg) was added to ropivacaine for three blocks in each group, which is known to accelerate the onset of regional anaesthesia.^16^ No dexamethasone, or other anaesthetic adjunct medication, was added to injectate for any patient. Injectate volume varied between groups, with providers mostly using 40mL (range 20-40mL) for FICB, 20mL (20-30mL) for FNB and 20mL (15-40mL) for PENG block.

### Outcome Measures

Pain scoring was scheduled prior to regional anaesthesia, at 15, 30 and 60 minutes post regional anaesthesia, and then hourly until 12 hours post regional anaesthesia or operative treatment. Clinicians asked enrolled patients to mark a 100mm Visual Analogue Scale (VAS) on the CRF (Supplementary material), otherwise clinicians used the Pain Assessment IN Advanced Dementia (PAINAD)^17^ tool for patients with severe cognitive impairment.

The VAS tool was selected to assess pain in patients without cognitive impairment as it has been widely used in the existing literature comparing FICB to FNB.^8,18,19^ PAINAD was also employed to facilitate the inclusion of patients with significant cognitive impairment, as it is superior to numerical rating scale (NRS) in this population.^20^ The protocol planned for VAS and PAINAD scores to be used interchangeably for analysis of the primary outcome, similarly to the NRS and PAINAD scores being used interchangeably in another study comparing FICB to FNB.^7^

Scores were documented by clinicians on the CRF on a scale from 0 to 10. When VAS was used, pain was scored at rest and on movement, defined as attempted 15 degrees of hip flexion. PAINAD scores were only recorded at rest, as the tool assesses behaviour over time. When pain scores were documented on the CRF but the VAS tool had not been marked, the investigators assumed that the NRS^21^ had been used, in keeping with routine practice at this centre. NRS has been validated to correlate with VAS.^21,22^

Opioid use was recorded pre-hospital, including by the ambulance service or at the referring hospital, and in the ED during the pre-block time period and until 12 hours post block or the patient entered the operating theatre. These data were available to the investigators via the electronic medical record and ambulance service patient records. Opioid use was converted to oral morphine milligram equivalent (MME) using the Australian and New Zealand College of Anaesthetists Faculty of Pain Medicine Opioid Calculator.^23^

Adverse events were identified from the electronic medical record, using truncated keyword search within the admission date range for the following terms: ‘aspiration’, ‘delirium’, ‘sedation’, ‘naloxone’, and ‘constipation’. The electronic medication chart was searched within the admission date range for naloxone and antibiotic prescription with medical records then reviewed to clarify any association with iatrogenic opioid overdose or infections potentially associated with regional anaesthesia.

The primary outcome was the maximal reduction in pain score at rest within 60 minutes of regional anaesthesia. Prespecified secondary outcomes included pain score reduction on movement, opioid use, onset time of regional anaesthesia (defined as the first recorded pain score of 1.5 less than the baseline score), and adverse events, including aspiration, delirium, sedation, constipation, or injection site infection. A post-hoc analysis of maximal pain score reduction for intracapsular versus extracapsular fractures was performed to assess any difference in the effectiveness of the PENG block. This is important as the innervation of the proximal femur distal to the joint capsule is not as clearly described as that of the anterior joint capsule.

### Sample size

The sample size calculation was based on previous literature, which used VAS,^8^ or PAINAD and NRS combined.^7^ We aimed to detect a clinically significant difference in pain score reduction of 1.5 out of 10 points with a standard deviation of 2.4, a power of 80% and a 2-sided alpha of .05. Based on previous literature,^7,19^ we assumed that the FICB and FNB groups would be sufficiently similar to combine as a single data set to compare to the PENG block, with a 2:1 allocation ratio. Therefore, the sample size calculated for the study was 74 patients, including at least 25 in the PENG block group. Based on an audit of proximal femur fractures at this centre, it was anticipated that enrolment of 80% of eligible patients over a 6 month period would achieve the target sample size. Consecutive patients were enrolled over 9 months from October 2019 to July 2020.

### Data Analysis

Data were entered into an Excel (version 2008, Microsoft, Redmond, WA) spreadsheet then analyzed in IBM SPSS Statistics (version 26; IBM). Normality of the data were assessed using the Shapiro-Wilk test. Basic descriptive statistics (counts, frequencies and mean and standard deviation (for normally distributed continuous variables) and median and interquartile range (IQR) for non-normally distributed data. Graphs of the primary outcome were produced in Python v37.0, showing actual data points, medians and the 95% confidence intervals around the medians.

## RESULTS

### Characteristics of study subjects

There were 67 eligible patients during the enrolment period, with 52 (78%) prospectively enrolled. Thirty-three received femoral blocks (19 FICB, 14 FNB) and 19 received a PENG block (Figure 2). Fifteen enrolments were missed by treating clinicians. Recruitment to the study was terminated early due to COVID-19 restrictions impacting the investigators’ ongoing access to the study site. All 52 eligible patients were included in the final analysis.

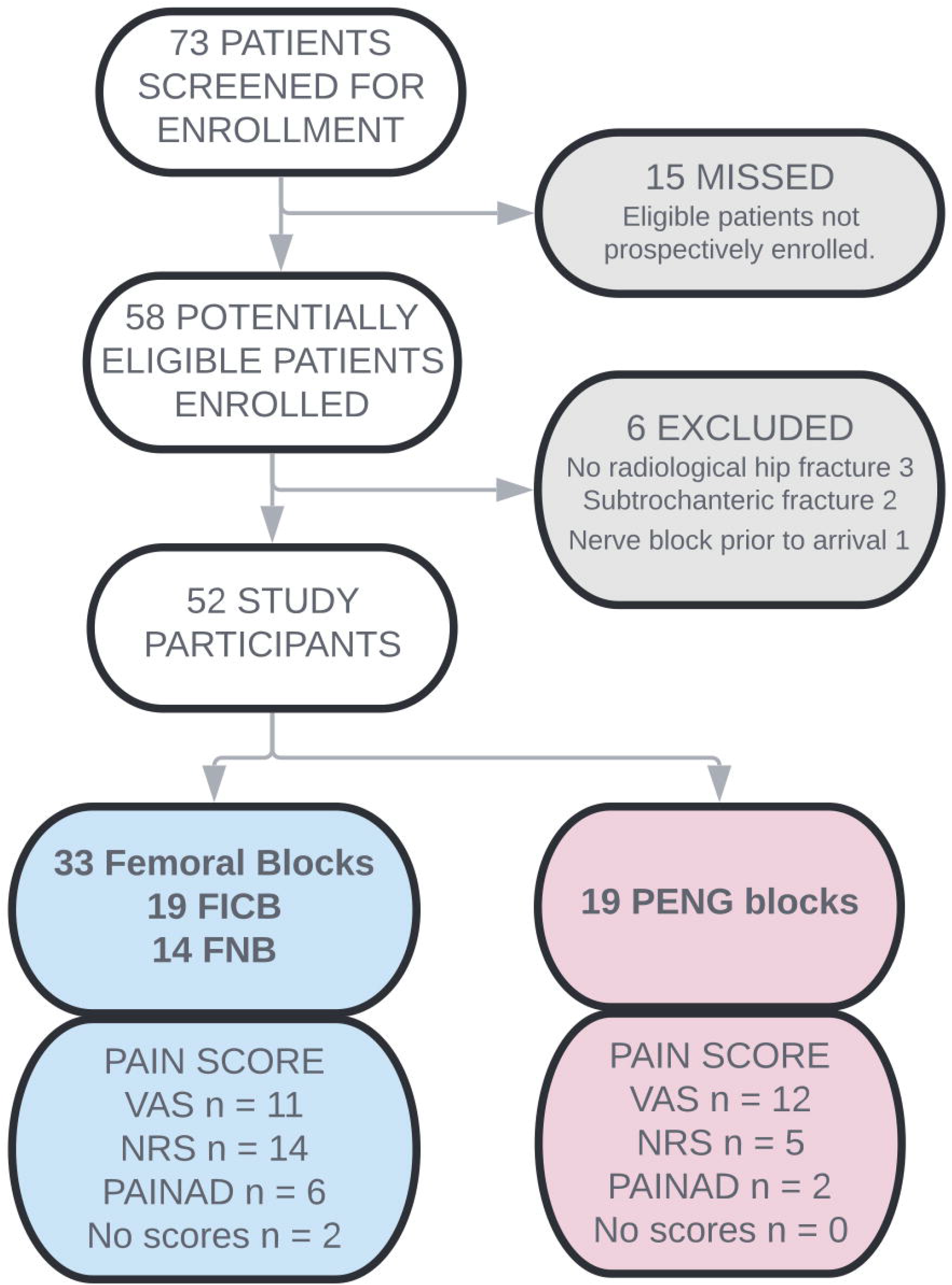

A comparative analysis was conducted of baseline characteristics of the two patient groups: FB and PENG block (Table 1). Patient demographics were similar between groups. Patients from the FB group were more likely to suffer chronic pain, although opioid use at baseline was similar. The groups differed in fracture type, with patients who received FB having a higher proportion of extracapsular fractures. Three patients had significant concurrent painful injuries, 2 from the PENG block group (contralateral Weber-C ankle fracture, anterior shoulder dislocation) and 1 from the FB group (distal radius fracture). Operators providing the PENG block were significantly less experienced in the technique than operators providing FB.

**Table 1.**
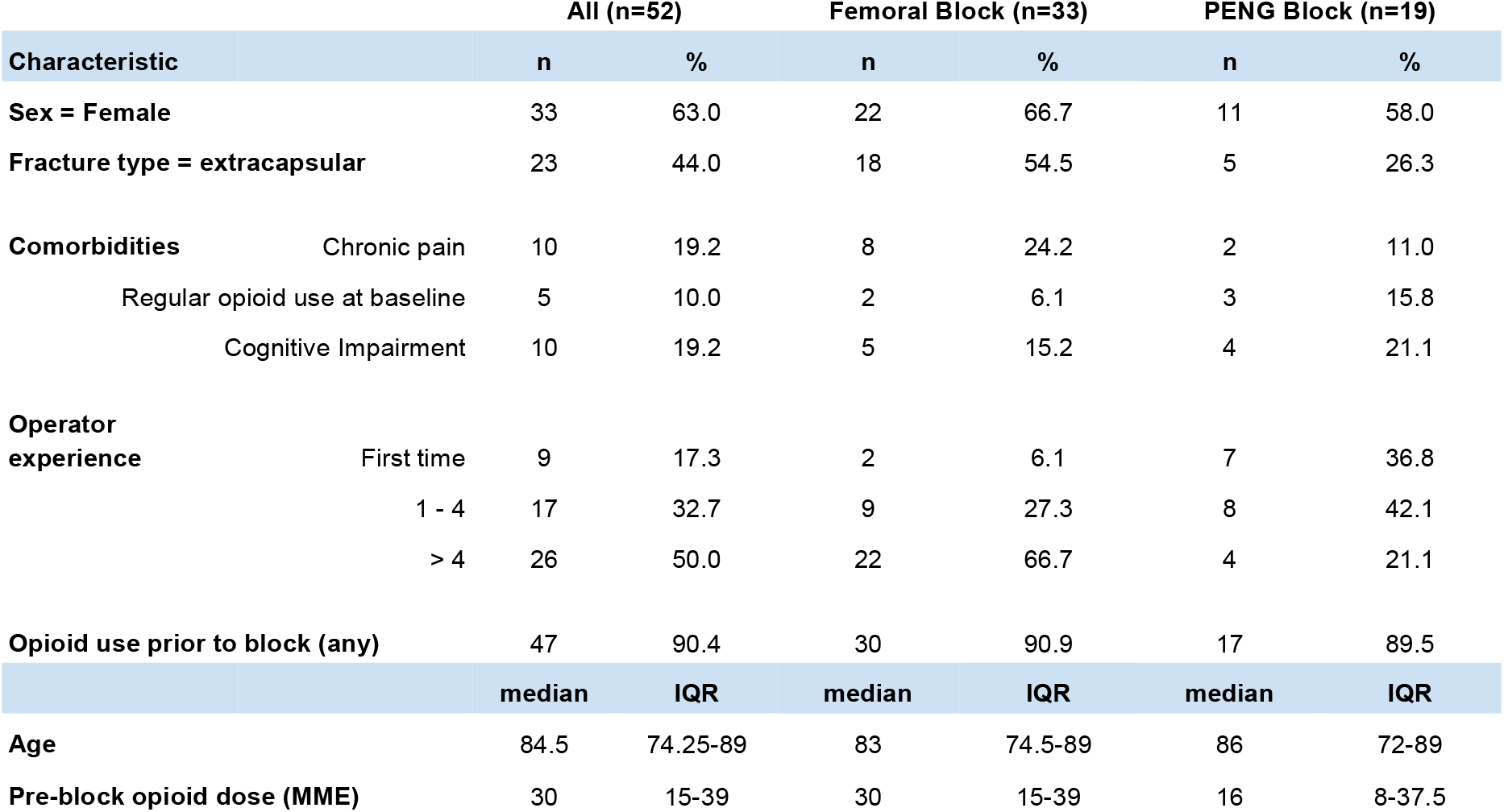
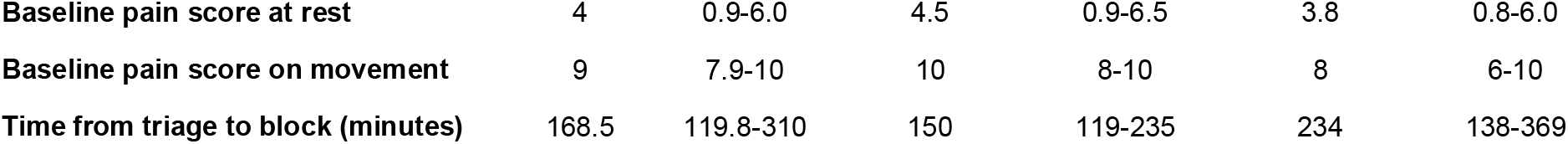
Baseline characteristics of study cohort. Femoral Block, fascia iliaca compartment block or femoral nerve block; PENG, pericapsular nerve group; MME, oral morphine milligram equivalent; IQR, interquartile range.

|  |  | All (n=52) |  | Femoral Block (n=33) |  | PENG Block (n=19) |  |
| --- | --- | --- | --- | --- | --- | --- | --- |
| Characteristic |  | n | % | n | % | n | % |
| Sex = Female |  | 33 | 63.0 | 22 | 66.7 | 11 | 58.0 |
| Fracture type = extracapsular |  | 23 | 44.0 | 18 | 54.5 | 5 | 26.3 |
| Comorbidities | Chronic pain | 10 | 19.2 | 8 | 24.2 | 2 | 11.0 |
|  | Regular opioid use at baseline | 5 | 10.0 | 2 | 6.1 | 3 | 15.8 |
|  | Cognitive Impairment | 10 | 19.2 | 5 | 15.2 | 4 | 21.1 |
| Operator experience | First time | 9 | 17.3 | 2 | 6.1 | 7 | 36.8 |
|  | 1 - 4 | 17 | 32.7 | 9 | 27.3 | 8 | 42.1 |
|  | > 4 | 26 | 50.0 | 22 | 66.7 | 4 | 21.1 |
| Opioid use prior to block (any) |  | 47 | 90.4 | 30 | 90.9 | 17 | 89.5 |
|  |  | median | IQR | median | IQR | median | IQR |
| Age |  | 84.5 | 74.25-89 | 83 | 74.5-89 | 86 | 72-89 |
| Pre-block opioid dose (MME) |  | 30 | 15-39 | 30 | 15-39 | 16 | 8-37.5 |

|  |  |  |  |  |  |  |
| --- | --- | --- | --- | --- | --- | --- |
| Baseline pain score at rest | 4 | 0.9-6.0 | 4.5 | 0.9-6.5 | 3.8 | 0.8-6.0 |
| Baseline pain score on movement | 9 | 7.9-10 | 10 | 8-10 | 8 | 6-10 |
| Time from triage to block (minutes) | 168.5 | 119.8-310 | 150 | 119-235 | 234 | 138-369 |

### Main results

There was no difference in pain score reduction within 60 minutes between groups when measured at rest or when measured on movement (Table 2). Absolute pain scores at 60 minutes were similar (Figure 3). Although 85% of the 162 scheduled pain scores were recorded in the first hour, only 3% of pain scores between 2-12 hours post block were recorded. No patient was lost to follow up.

**Table 2.** Outcomes by block type. Femoral Block, fascia iliaca compartment block or femoral nerve block; PENG, pericapsular nerve group; MME, oral morphine milligram equivalent; IQR, interquartile range.

| Characteristic | All (n=52) |  | Femoral Block (n=33) |  | PENG block (n=19) |  |
| --- | --- | --- | --- | --- | --- | --- |
| Scale and continuous variables | Valid n | Median (IQR) | n | % | Valid n | Median (IQR) |
| Maximum pain score reduction at rest | 50 | 2 (0 - 5.13) | 31 | 2 (0-6) | 19 | 2 (0-5) |
| Maximum pain score reduction on movement | 40 | 3.2 (2.0-5.0) | 24 | 3.2 (2.0-5.0) | 16 | 3.4 (1.0-5.1) |
| Time of onset assessed at rest (minutes) | 28 | 15 (15-30) | 17 | 15 (15-60) | 11 | 15 (15-30) |
| Time of onset assessed on movement (minutes) | 32 | 15 (15-30) | 21 | 30 (15-30) | 11 | 15 (15-30) |
| Post block opioid use 6 hours post block (MME) | 52 | 7.5 (0-15) | 33 | 7.5 (0-15) | 19 | 0 (0-10) |
| Post block opioid use 12 hours post block (MME) | 52 | 21.25 (7.5-37.5) | 33 | 23 (15-37.5) | 19 | 8 (0-25.75) |
| Total opioid use at 6 hours (MME) | 52 | 32 (21-46.5) | 33 | 37.5 (22.5-52) | 19 | 30 (8-37.5) |
| Total opioid use at 12 hours (MME) | 52 | 44.25 (30.0-68.25) | 33 | 52.5 (31-82.5) | 19 | 39 (8-45.5) |
| Duration of admission (days) | 52 | 9 (6-11.75) | 33 | 8 (6-10) | 19 | 10 (6-15) |
| Yes or no variables | Valid n | n (%) | Valid n | n (%) | Valid n | n (%) |
| Opioid free for 6 hours post block | 52 | 21 (40.4) | 33 | 11 (33.3) | 19 | 10 (52.6) |
| Opioid free for 12 hours post block | 52 | 9 (17.3) | 33 | 3 (9.1) | 19 | 6 (31.6) |
| Rescue block during first 6 hours | 52 | 2 (3.8) | 33 | 1 (3.0) | 19 | 1 (5.3) |
| New onset delirium during admission | 52 | 11 (21.2) | 33 | 9 (27.3) | 19 | 2 (10.5) |
| New onset constipation during admission | 52 | 9 (17.3) | 33 | 7 (21.2) | 19 | 2 (10.5) |

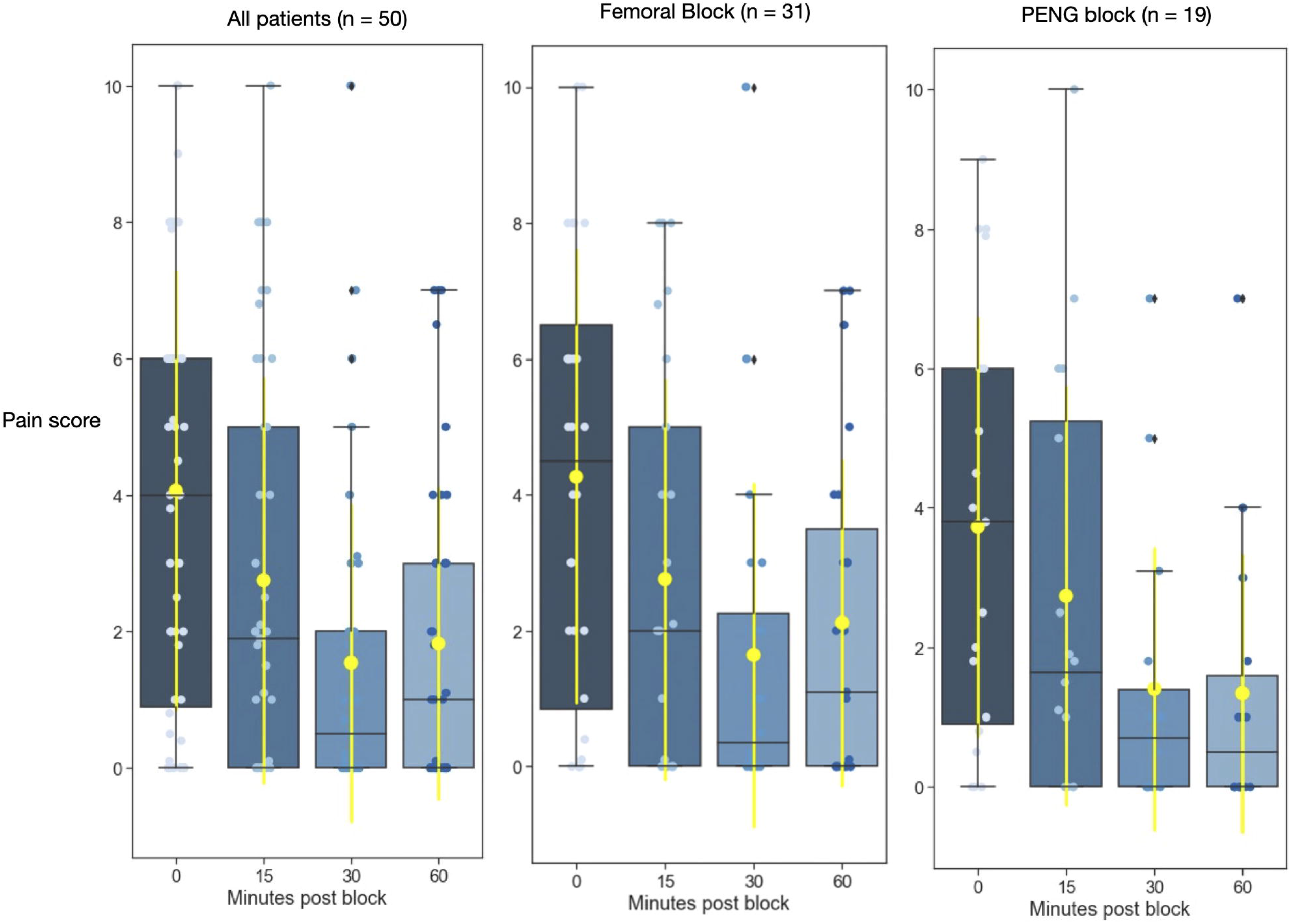

Opioid use was statistically similar in the 12 hour period after regional anaesthesia (median MME of 8 mg for PENG block vs 23 mg in the FB group (Table 2). However, more patients were opioid free for 6 hours after regional anaesthesia in the PENG block group (53% vs 33% FB) and for the duration of follow up (31% vs 9% FB). The median onset times were similar.

Adverse events were similar between groups (Table 2). No regional anaesthesia associated infections were detected during admission. One patient from each group required a rescue block provided by an anaesthetist prior to operative fixation. One patient with a FICB had a repeat FICB using landmark technique. One patient with a PENG block had a femoral nerve catheter inserted. Three patients were checked in to the operating theatre prior to twelve hours post regional anaesthesia (1 PENG block, 2 FB).

The PENG block appears as effective for extracapsular proximal femur fractures compared to intracapsular fractures. (Figure 4). As this is the first study to compare the PENG block to FICB or FNB in the ED, or any preoperative setting, demographics (supplementary table 1) and outcome data (supplementary table 2) are provided as three patient groups (FICB, FNB, PENG block) and raw data (supplemental material) is provided to facilitate future meta-analysis.

## DISCUSSION

In this pragmatic, single centre observational study of patients with hip fracture in ED, there was no difference observed in maximal pain score reduction within 60 minutes for the PENG block compared to FB. However, this study did demonstrate that the PENG block was feasible, safe and effective when provided in the ED by a group of providers relatively inexperienced in the technique.

Prior to this study, literature supporting the PENG block for hip fractures has been limited.^15^ There have been three case series and two case reports, amounting to a total of 22 patients, on the use of PENG block for effective analgesia of hip fractures.^15^ Existing comparative literature for the PENG block consists of a single cohort study of a convenience sample of 42 perioperative patients that compared postoperative opioid requirement after PENG block or FNB and demonstrated no difference.^10^ This study is the first to report on provision of the PENG block as part of routine ED practice and the first study to prospectively compare the PENG block to other techniques (FICB and FNB) for analgesia of hip fracture in the ED.

The PENG block has several characteristics highlighted by this study that make it attractive to ED clinicians. Firstly, it was straightforward for inexperienced providers to perform successfully, which is likely due to bony sonographic landmarks and tactile bony endpoint of needle insertion. Also, it is safe to perform as the target injection site is distant from vascular structures, innately minimising risk of local anaesthetic systemic toxicity, and there were no reported differences in adverse events compared to FB.

Opioid-sparing is important in the elderly population due to its potential morbidity. In our study, more patients were opioid free after a PENG block, adding weight to the potential for routine use in this setting. The data on opioid use was robust, with excellent capture due to the strict mandatory recording of these regulated medications. Additionally, we were able to account for potential confounders between groups, including concurrent injuries, baseline opioid tolerance or attending the operating theatre less than 12 hours from regional anaesthesia.

The effect of the PENG block for extracapsular versus intracapsular injuries has not been previously described. In terms of the proposed mechanism of action, the innervation of the extracapsular proximal femur is not yet as clearly delineated as the anterior hip capsule. Although the sample size was too small to draw any firm conclusion, the PENG block appears to be highly effective for extracapsular fractures (Figure 4). This limited data suggests that the extracapsular proximal femur receives nociceptive innervation from nerves that traverse the iliopsoas plane, and are anaesthetised by the PENG block.

A strength of this study was that the protocol prescribed use of VAS and PAINAD to allow inclusion of patients with cognitive impairment, who are often excluded from regional anaesthesia research^24^ despite representing a third of the disease burden for hip fracture.^19,24^ Having clinicians, rather than research assistants, enrol patients allowed for a 78% prospective recruitment rate with consecutive (24/7) screening. However, relying on the treating team to document pain scores resulted in NRS being used in place of VAS in 19 of 44 patients. Fortunately, NRS reduction has been validated to correlate with VAS reduction,^21,22^ so these scores were pragmatically included in our study.

Combining FICB and FNB as a single treatment arm for data analysis was reasonable based on previously established similar effectiveness.^7,19^ However, when our data was divided into three treatment arms, we found the FICB and FNB group differed significantly in baseline pain scores and pain score reduction was not equal. Due to early termination of recruitment, this study was potentially underpowered to identify any difference in the effectiveness of the PENG block compared to FICB and FNB.

There were several other limitations to this study. Prospective registration of patients mitigated recall bias, although pain scores were poorly recorded after the first hour. Given the open-label nature of the study, clinicians were aware of the intervention of interest and could have been subject to the Hawthorne effect.^25^ Although the PENG block is purported to be motor sparing, we did not report motor function in this study as it is more relevant in the postoperative period when lower limb weakness may delay safe mobilisation. Pain score recording after 60 minutes was too sparse to provide any information on the duration of action of any of the regional anaesthetic techniques provided. Adverse events were recorded retrospectively, although all the important adverse events were likely to have been captured.

This novel comparative study of the PENG block versus FB for proximal femur fracture in the ED provides important data on the feasibility, safety and effectiveness of the block. A larger multi-centre randomised controlled study should now be designed to determine whether the PENG block is superior to FICB or FNB for patient centred outcomes and to determine if any difference exists for different hip fracture subtypes.

## Data Availability

All data referred to in the manuscript will be publically available

## Acknowledgements

This trial was made possible with support of the Northern NSW Health Service and Lismore Base Hospital. We thank Justin Dryden for conducting the retrospective analysis preceding this study, and for data collection and entry. We thank Neil Stokes, Gerben Keijzers, Alexandre Stephens for their guidance in moving this study from an idea to a reality, and Morwenna Haywood for Figure 1A. We thank Stefan Jantschulev for his enthusiasm for the project and teaching us the PENG block. Abridged results were presented as a poster at the Australia New Zealand College of Anaesthesia (ANZCA) special interest group rural practice meeting on 13 June 2021 at Airlie Beach, Queensland, Australia. Awarded Mackay Institute of Research and Innovation Rolling trophy.

